# crossNN: an explainable framework for cross-platform DNA methylation-based classification of cancer

**DOI:** 10.1101/2024.01.22.24301523

**Authors:** Dongsheng Yuan, Robin Jugas, Petra Pokorna, Jaroslav Sterba, Ondrej Slaby, Simone Schmid, Christin Siewert, Brendan Osberg, David Capper, Pia Zeiner, Katharina Weber, Patrick Harter, Nabil Jabareen, Sebastian Mackowiak, Naveed Ishaque, Roland Eils, Sören Lukassen, Philipp Euskirchen

## Abstract

DNA methylation-based classification of brain tumors has emerged as a powerful and indispensable diagnostic technique. Initial implementations have used methylation microarrays for data generation, but different sequencing approaches are increasingly used. Most current classifiers, however, rely on a fixed methylation feature space, rendering them incompatible with other platforms, especially different flavors of DNA sequencing. Here, we describe crossNN, a neural network-based machine learning framework which can accurately classify tumor entities using DNA methylation profiles obtained from different platforms and with different epigenome coverage and sequencing depth. It outperforms other deep- and shallow machine learning models with respect to precision as well as simplicity and computational requirements while still being fully explainable. Validation in a large cohort of >1,900 tumors profiled using different microarray and sequencing platforms, including low-pass nanopore and targeted bisulfite sequencing, demonstrates the robustness and scalability of the model.

## Introduction

DNA methylation plays an important role in regulation of gene expression and cell type differentiation^1,2^. Patterns of 5-methylcytosine (5mC) define physiological cell states, but have also been linked to many human diseases, including cancer^3-5^. In medicine, epigenome-wide patterns of 5mC can be exploited for disease classification^6^. In particular, DNA methylation-based classification of tumours has emerged as a powerful conceptual and diagnostic tool both for establishing a clinical diagnosis and for investigating the molecular taxonomy of cancer^7-9^. Indeed, classification of central nervous system tumors has been embraced by the World Health Organization (WHO)^10^ with profound impact on routine diagnostic workup^4,5^. Moreover, integrated, histo-molecular classification of brain tumours hass been shown to refine histological diagnosis with reclassification in about 12% of cases^8^. Most implementations of diagnostic assays rely on generation of methylation profiles by hybridization microarray and supervised classification against a well-annotated reference set^11,12^ which has become a widely accepted diagnostic approach in adult and pediatric neuro-oncology^8,13-15^.

However, various methods for probing the 5mC methylome have been developed and benchmarked, each providing information on DNA methylation in different target regions and at different levels of resolution^16^. For example, whole genome bisulfite sequencing (WGBS) has long been seen as a gold standard in providing the most comprehensive DNA methylation map at single base resolution^17^. WGBS is expensive, however, and demands significant quantities of input DNA. Moreover, the sequenced reads often lack useful methylation information^18^. Targeted methylation sequencing (targeted methyl-seq) using restriction enzymes or, more recently, hybridization capture for enrichment has gained widespread popularity for cost-efficient targeted capture^19,20^. Microarray-based technologies, such as Infinium HumanMethylation450 (Infinium 450K) and Infinium HumanMethylation850 (MethylationEPIC, EPIC) also have been widely employed to survey specific genomic loci across the genome with bespoke probes^21^. More recently, third generation sequencing techniques have allowed base modifications from natural DNA to be inferred. We and others have demonstrated suitability and robustness of low-coverage whole-genome nanopore sequencing in clinical application for accurate, rapid, and cost-efficient DNA methylation-based classification of brain tumours^22,23^. However, the commonly aimed for ultra-low sequencing depth and coverage leads to mostly binary methylation information (instead of beta values) of a random subset of the ∽30 million CpG sites in the genome^23^.

All these methods have been found deliver highly concordant results, but different genomic coverage and depth have so far required different classification assay-specific approaches^24^. Various machine learning algorithms have been used for the task of DNA methylation-based classification but are mainly restricted in single platform data or fixed feature spaces, e.g. the most commonly used random forest (RF) model for use with microarray data^8^. Previously, we proposed ad-hoc RF which can bridge the gap between low-coverage nanopore sequencing data and microarray reference data at the expense of training an ad-hoc new model for each unknown sample which, however, is time-consuming, computationally expensive and introduces non-comparability between these patient-specific models^23^. Recently, a neural network-based model has been proposed using sparse data to predict brain tumor classes^25^. A precise model that can predict brain tumour classes across platforms is still urgently needed.

Here, we propose crossNN, a unified neural network-based framework trained on fixed reference data that handles variable and sparse feature sets for prediction. The model enables instantaneous predictions from methylation profiles generated by multiple platforms including WGBS, targeted methyl-seq, low-coverage nanopore WGS and various microarray platforms (Illumina 450K, EPIC, EPICv2). At the same time, the lightweight scalable architecture allows for rapid re-training and cross-validation for the rapidly emerging landscape of cancer reference atlases.

## Results

### Model development and workflow

The crossNN model architecture (Figure 1) relies on a perceptron, implemented as single layer neuronal network using pytorch (Online Methods). The network architecture consists of only input layer and output layer with the two layers being fully connected without bias, which means the model will capture the linear relation between the input CpG sites and methylation classes. For training, we used the Heidelberg brain tumour classifier v11b4 reference set comprising methylation profiles of 2801 samples from 82 tumour types and subtypes (methylation classes, MC) and 9 non-tumor control classes, generated using Illumina 450K microarrays^8^. The feature space of the training set is fixed given the array probe set and mainly covers CpG sites in CpG islands and promoter regions.

**Figure 1:**
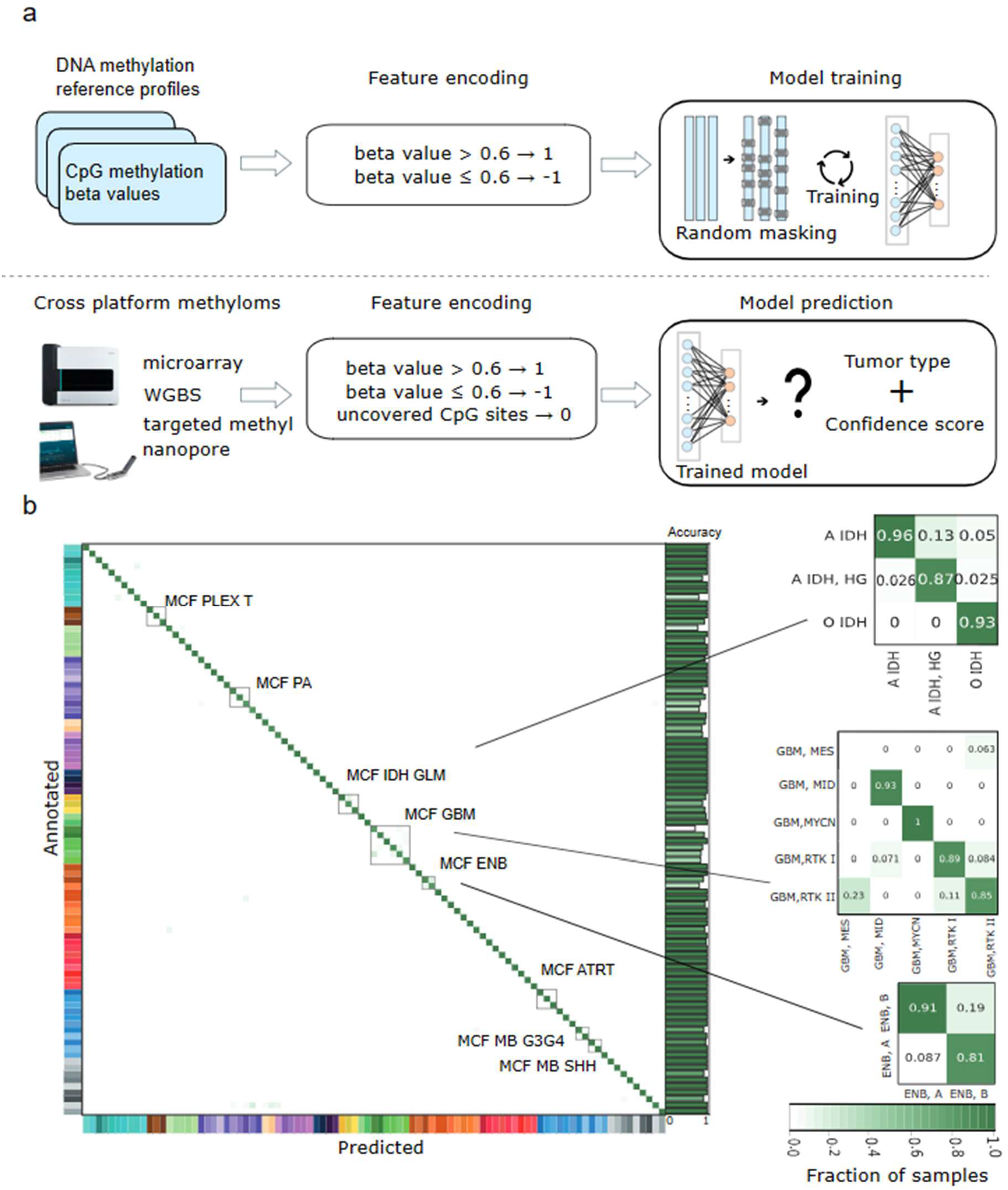
(a) Overview of model the model architecture. (b) Heatmap of confusion matrix in 5-fold cross-validation. WGBS, whole genome bisulfite sequencing. MCF, methylation class family. Abbreviations of methylation classes are in line with the original publication of the training set by Capper et al.

During pre-processing and for cross-platform normalization, CpG sites in the training set were binarized using an empirically determined beta value threshold of 0.6^23^. Thereafter, uninformative probes were removed (see *Online Methods*), resulting in a total of 366,263 binary features.

To enable tumor classification using different platforms for methylome profiling with varying or sparse epigenome coverage, the model was trained with randomly and repeatedly masked input data. The masked CpG sites during training were encoded as zero, unmethylated sites as -1 and methylated probes as 1. The model was then trained using the randomly resampled and [-1,1]-encoded binary training set. For prediction from methylation profiles from different platforms, methylated allele frequencies at CpG sites were equally binarized and missing features encoded as zero.

Critical hyperparameters that were optimized included masking rate p and number of epochs e (which is proportional to how many times each sample is resampled). Using a grid search approach, a masking rate of 97.5% and e = 1000 epochs were selected for training the final model (Supplementary Figure 1).

### Evaluation of model performance

First, model performance was validated by 5-fold cross-validation (CV) in the training dataset. Overall accuracy was 96.11 ± 0.86 % across all CV at methylation class (MC) level (Supplementary Figure 2). Tumor classes within the same methylation class family (MCF) are closely and misclassifications inside MCF will usually not have clinical impact. Indeed, most misclassifications were observed within MCF (Figure 1b). Therefore, at MCF level, prediction accuracy reached 99.07 ± 0.21%. In comparison, ad hoc random forest models for each subsampled feature set reached lower accuracy both at MC level and MCF level (94.93 ± 0.88% and 97.89 ± 0.60%, respectively).

To further test our model’s performance with samples with different coverages of the CpG sites, the microarray samples on the test folds were sub-sampled with different sampling rates from 0.5% to 100% and for each sample rate we repeated this process randomly 10 times. Our model showed robust performance with high average accuracy in 5-fold cross-validation with different sampling rates from 0.5% to 75% (Supplementary Figure 2).

### Independent cross-validation in different platforms

Next, we validated the final model in independent cohorts generated on different microarray and sequencing platforms. We assembled a validation cohort totalling 1,923 patient samples generated on Illumina 450K (N=610), EPIC (N=649) and EPICv2 (N=10) microarrays as well as nanopore low-pass WGS (N=415), Illumina targeted methyl-seq (N=124) and Illumina WGBS (N=125) sequencing (Supplementary Table 1). The validation set covered 65 different brain tumor types, reflecting 72 out of the 81 methylation classes in the training set.

**Table 1:**
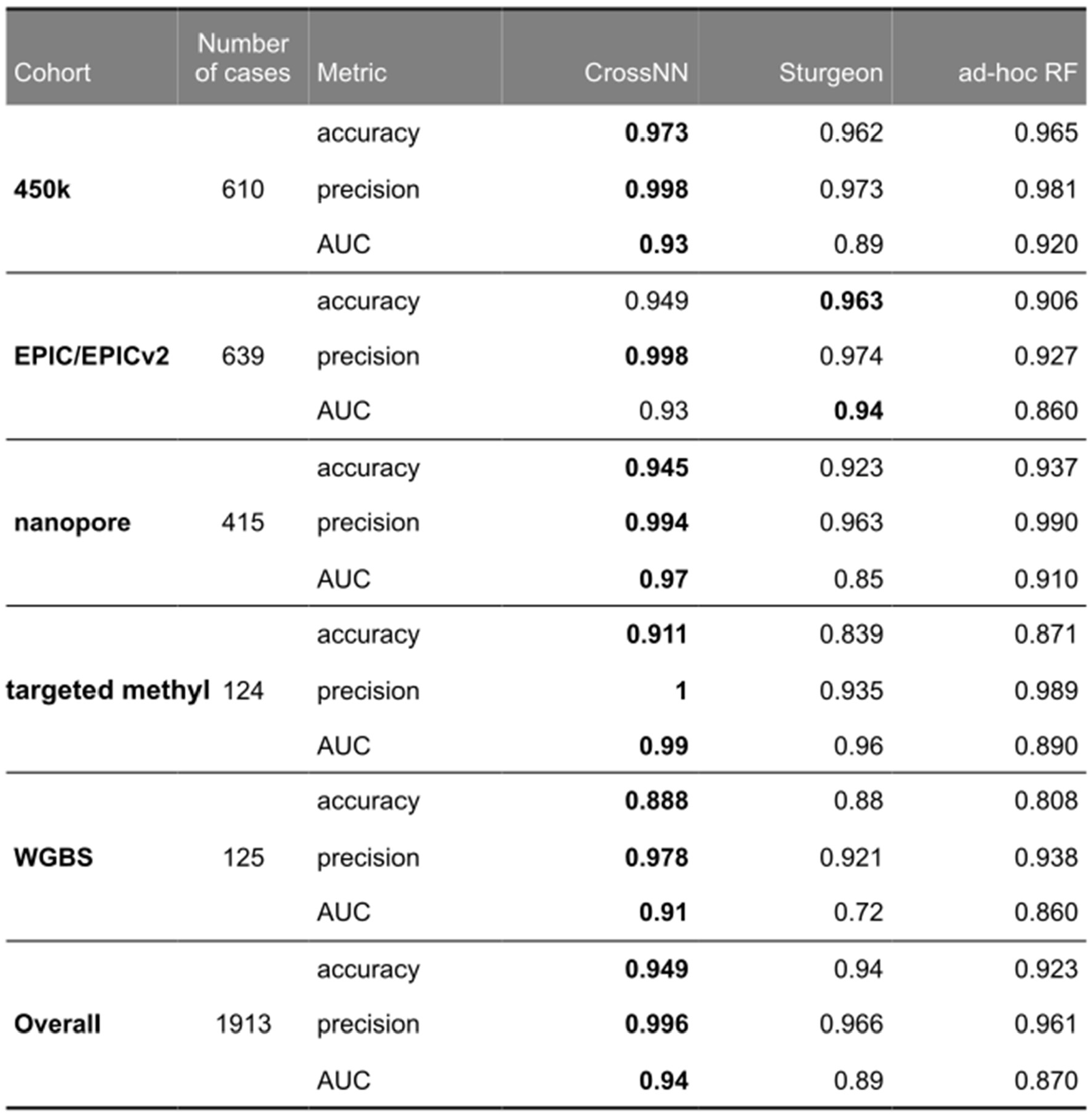
Comparison of the crossNN model to ad-hoc random forests (^23^) and the “Sturgeon” deep neural network approach (^25^). For each model, raw accuracy before application of cut-offs, precision with platform-specific cut-offs and area under the curve of the ROC curve for the (calibrated) score to predict correct classification are given. For crossNN, the following cut-offs as derived above are used: microarray > 0.4; crossNN nanopore/targeted methyl-seq/WGBS > 0.2. Published validated cut-offs were used for ad-hoc RF and the sturgeon DNN (ad hoc RF > 0.15; sturgeon DNN > 0.8). *AUC, area under the curve. ROC, receiver-operator characteristics. DNN, deep neural network. NN, neural network. RF, random forest. WGBS, whole genome bisulfite sequencing*.

Depending on the assay, the distribution of the number of CpG features used for prediction varied by two orders of magnitude (Fig. 2a). Nevertheless, we achieved a high overall accuracy of 0.95 and AUC of 0.94 (ranging from 0.93 to 0.99 per platform, Fig. 2c). Because the distribution of scores of predictions made by the model varied across platforms (Fig. 2b), we identified platform-specific diagnostic cut-offs for correct classification using per-platform 5-fold CV. The optimal cut-off in each fold was determined using the Youden index in the ROC curves (Supplementary Figure 3). The range of optimal cut-offs was similar for microarray vs. sequencing platforms (Fig. 2b, indicated in *blue*). For simplicity, we therefore conservatively selected a cut-off > 0.4 for all microarray platforms and > 0.2 for all sequencing platforms. This resulted in precision > 0.97 for all platforms.

**Figure 2:**
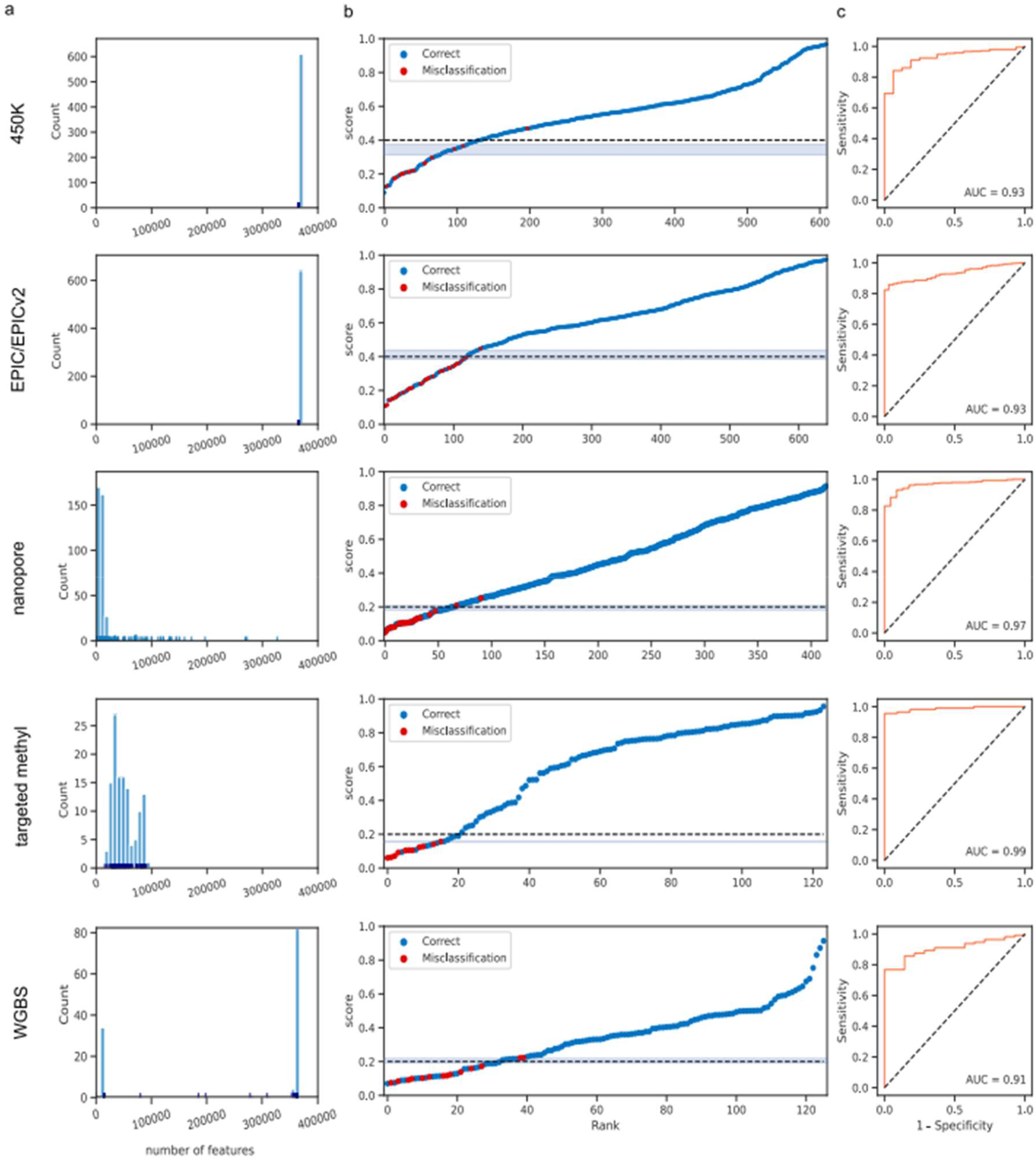
Classification result in 450K, EPIC/EPICv2, nanopore, targeted methyl-seq, WGBS cohorts. (a) Distribution of the number of CpG features used for prediction. (b) Scatter plot of cohorts with samples ranked by confidence score. Dashed lines were showing the cut-off values. (c) Receiver operator characteristics of the score to predict correct classification per platform. *AUC*, area under the curve.

### Comparison to other algorithms

Next, we compared model and cut-off performance to our previously published ad-hoc random forest (*ad hoc RF*) approach^23^ and a recently published deep-neural network (*Sturgeon DNN*)^25^. All approaches have been developed to make predictions from sparse nanopore data, yet can be applied to any source of methylation data and use an identical training set.

Our shallow neural network model was non-inferior to ad-hoc RF and the Sturgeon DNN with respect to overall accuracy and outperformed both approaches in terms of ROC characteristics of the prediction scores, especially precision (Table 1).

### Interpretability of the model

Finally, our model’s architecture facilitates interpretability by capturing the linear relationships between CpG probes and tumor classes or sub-classes. The weights of the edges connecting the input CpG features and the output layer thus can be interpreted as indicators of feature importance for each tumor (sub)type. These weights offer insights into the significance or relevance of individual CpG probes in the classification of specific tumor types: Each CpG feature will be assigned a positive or negative weight for each tumor type. Positive weights indicate that if a given CpG site is methylated, the sample is more likely to match the corresponding tumor type, and vice versa.

The absolute value of the weight reflects the importance of a given CpG site in predicting the associated tumor type. For each tumor type, CpG sites with top positive/negative weights are differentially methylated between tumor (sub)types, which can be helpful to reveal biological mechanisms underlying tumor type identity such as cell of origin and discover potential biomarkers (Figure 4a,b).

**Figure 4:**
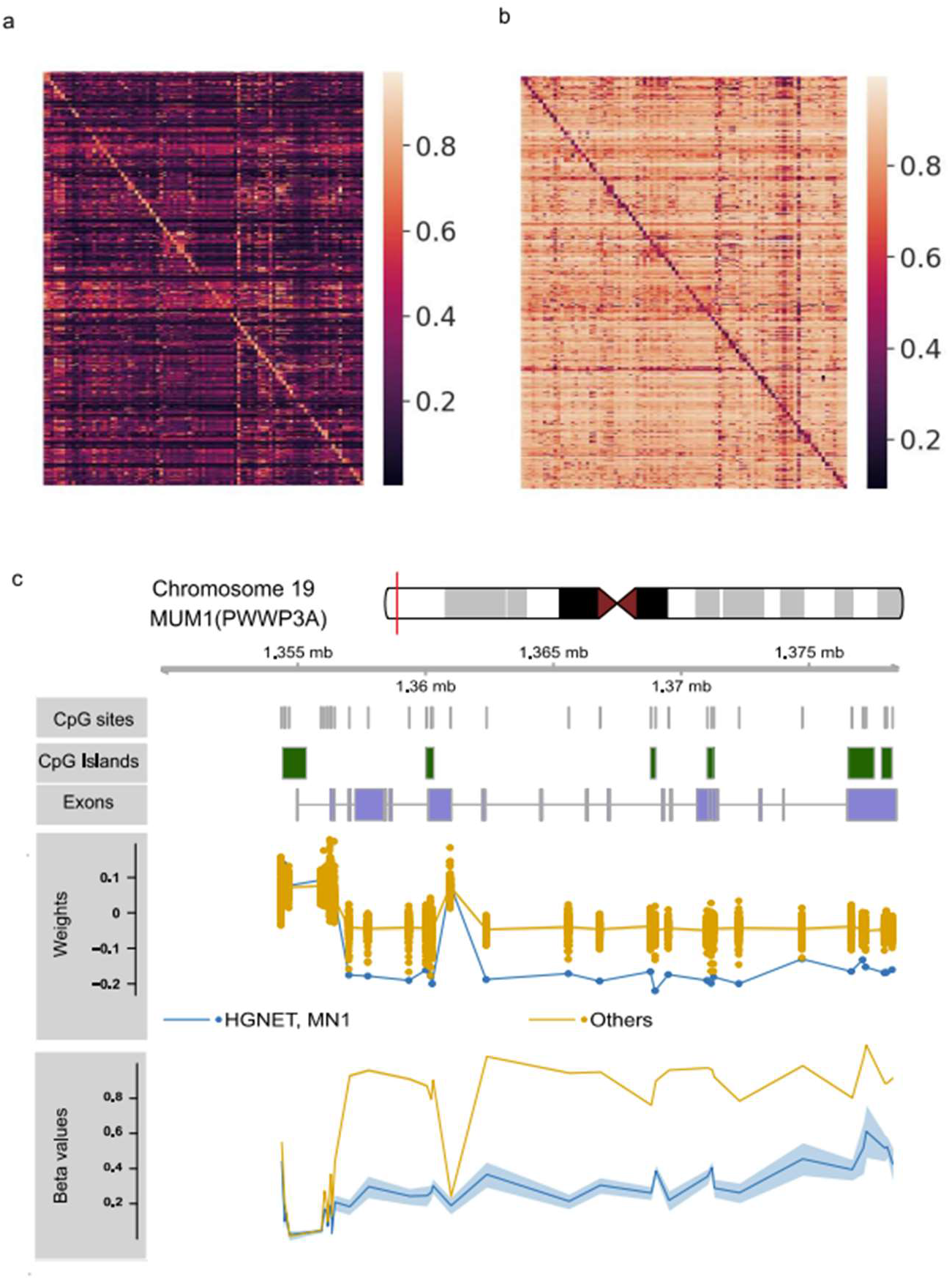
Interpretatibility of the model. (*a,b*) Heatmaps demonstrate methylation levels (beta value) of the top 100 CpG sites associated with a given tumor type, ranked by feature weight in the final prediction model. Features with positive (*a*) and negative (*b*) weights were ranked independently. (*c*) Importance of class-specific features with respect to genomic context. PWWP3A gene which was identified as marker gene for the methylation class high grade neuroepithelial tumors with MN1 alterations (HGNET, MN1) using ranking of feature weights aggregated on gene level. Differential hypomethylation is observed in the gene body, but not a proximal CpG island (lower track).

For example, CpG sites within the *PWWP3A* gene locus were ranked most important for prediction of the methylation class *high-grade neuroepithelial tumor with MN1 alteration* (HGNET-MN1) a novel tumor type which has recently been endorsed by the WHO 2021 classification as astroblastoma, MN1-altered^10^. In accordance with the negative weight of most features, the PWWP3A gene body was hypomethylated (Fig. 4C, lower track). Indeed, mRNA expression of *PWWP3A* has previously been identified as marker gene for HGNET-MN1^26^. Many CpG sites in the gene body of *PWWP3A* show remarkable negative weights in our model to HGNET-MN1 subtype comparing to other tumor types while the CpG island upstream the transcription start site is not informative for discriminating HGNET-MN1 (Fig. 4c). Thus, feature importance revealed by the model sheds light on functional importance of individual (marker) genes and hints at positional importance of epigenetic modifications within a gene’s structure.

## Discussion

In this study, we present a novel and simple machine learning framework which can accurately classify tumor entity using DNA methylation profiles obtained from different platforms and with different epigenome coverage and sequencing depth. It outperforms other deep- and shallow machine learning models with respect to precision as well as simplicity, computational requirements (for both training and prediction) while still being fully explainable. Validation in low-pass nanopore WGS, WGBS, targeted methyl-seq and microarray brain tumor cohorts demonstrates the robustness and scalability of the model.

Mainly developed for sparse methylomes generated by ultra low-pass nanopore WGS, this pretrained model enables predictions within seconds, outperforming our previous ad hoc random forest implementation which required time-consuming and computationally intense re-training for individual samples^23,27^. Immediate predictions greatly improve time-critical applications such as intraoperative diagnostics. In comparison to a recently published deep neural network model^25^ trained on the same dataset, it performs non-inferior with respect to overall accuracy and is superior with respect to precision when applying diagnostic cut-offs on prediction scores which is critical to ensure high specificity in clinical application. At the same time, the light-weight architecture allows rapid training on novel reference sets.

Despite using a neural network architecture, the model maintains a simple linear structure, which limits overfitting and drastically increases the interpretability of the model. Feature importance guides biological and clinical interpretation of the model and facilitates marker gene detection in each tumor type.

Importantly, the model is compatible with the EPICv2 microarray platform whose probe set is not downward-compatible and precludes use of most versions of the original Heidelberg brain tumour classifier. We provide an intuitive web-based graphical user interface that allows users to upload methylation data and predict tumor entity instantaneously (https://crossnn.dkfz.de). Additionally, models and source code are available for local deployment and integration with institutional workflows (https://gitlab.com/euskirchen-lab/crossnn).

### Limitations

We employed binarization of methylated allele frequencies as a means for cross-platform normalization and feature encoding. However, using an empirically chosen global cut-off for binarization might be sub-optimal for some methylation classes and introduce bias. For tumor types with global hypo-or hypermethylation (such as pituitary tumors or IDH-mutant glioma, respectively) or low tumor purity due to complex tumor microenvironment, such as mesenchymal subtype IDH-wildtype glioblastoma^28^, it might introduce a class-specific bias which remains to be investigated systematically.

Despite a large validation cohort (>1,900 patients) in this study, rare brain tumour types were under-represented or omitted. Thus, ongoing validation in very large multicentric cohorts covering the full spectrum of brain tumours using different techniques are warranted to fully characterize class-specific model performance and identity potential bias.

In conclusion, our study offers a machine learning framework for cross-platform DNA methylation-based classification of cancer, enabling development of rapid, resilient, interpretable, and accurate diagnostic tests. These methods hold promise to become valuable diagnostic tools for all types of cancer well beyond neuro-oncology.

## Online Methods

### Public datasets

The reference set of the Heidelberg brain tumour classifier v11b4 (GSE90496) containing 2,801 samples and 82 types of brain tumors and 9 control classes was used for model training^8^. For validation, preprocessed public datasets from the following studies were integrated from the sources indicated: medulloblastoma WGBS^29^ downloaded from the IGCG Data portal (https://dcc.icgc.org/releases/release_28/Projects/PBCA-DE), GSE121721 for glioblastoma WGBS^30^, GSE209865 for nanopore low-pass WGS ^23^, GSE109379 for 450K microarray^8^.

### Methylation microarrays

DNA methylation and copy number analyses were performed using the Infinium Methylation450k and EPIC Bead-Chip array platforms (Illumina, USA). All analyses were performed according to the manufacturer’s instructions. In brief, DNA was extracted from FFPE tumor samples using the Maxwell RSC FFPE Plus DNA Purification Kit (Promega, USA). After bisulfite conversion using the Zymo EZ Methylation Kit (Zymo Research Irvine, USA), the Infinium HD FFPE DNA Restore Kit was used for DNA restoration. The beadchips were scanned on the iScan system (Illumina, USA). The unprocessed output data (.idat files) from the iScan reader were checked for general quality measures as indicated by the manufacturer.

### WGBS sequencing and processing

Libraries were prepared using a NEBNext Methyl-seq Kit (NEB), and were then sequenced on an Illumina NovaSeq 6000 platform (instrument A01077) at the BIH Core Unit Genomics over two S4 flow cells in a paired-end setting of 2 × 150 bp. Processing of WGBS data from 22 human diffuse glioma samples was performed using the One Touch Pipeline (OTP)^31^ which uses bwa v0.6.1^32^ for alignment and methylCtools v1.0.0^29^ for methylation calling. Plus- and minus-strand methylated allele frequencies at CpG sites were merged using custom scripts. The mean mapping rate was 99.96% (range 99.93-99.9899%) with 95.7% properly paired (range 91.2-98.1%) and a 10.2% duplication rate (7.6-13.8%). Alignment resulted in a mean coverage of 70.5×4× per sample (range 57-89x),

### Targeted methylation sequencing and processing

Frozen tumor tissues collected during surgery aiming for partial or total tumor resection was used as source material for DNA extraction, which was performed using mechanic homogenization with ceramic beads and subsequent column-based extraction with DNeasy Blood & Tissue Kit (Qiagen). Prior to library preparation, DNA was quantified using Qubit dsDNA BR Assay Kit (Invitrogen). Sequencing libraries were prepared either with TruSeq-Methyl Capture EPIC Library Prep Kit (Illumina) or a combination of SureSelectXT Methyl-Seq Library Preparation Kit with SureSelectXT Human Methyl-Seq target enrichment panel (Agilent). Sequencing libraries prepared with TruSeq-Methyl Capture EPIC Library Prep Kit were sequenced on the NextSeq 500 device using NextSeq 500/550 Mid Output Kit v2.5 (150 cycles) (Illumina) in a paired-end setting of 2 × 80bp. Libraries prepared with SureSelectXT Methyl-Seq panel were also sequenced on the NextSeq 500 device using either NextSeq 500/550 Mid Output Kit v2.5 (300 cycles) or NextSeq 500/550 Mid Output Kit v2.5 (150 cycles) in a paired-end setting of 2 × 151bp and 2 × 80bp, respectively. Sequencing reads were quality checked with FastQC v0.11.9^33^. Adapters and low-quality 3’ ends trimming was done with TrimGalore^34^. The alignment to human reference hg19 and methylation calling were carried out completely with Bismark v0.23.1^35^.

### Feature selection

First, probes that were always methylated or un-methylated across all the samples were considered as uninformative and were removed from the dataset.

In the feature processing step, to fill the gap of different sequencing depths, all the methylated probes were encoded as 1 and correspondingly the unmethylated probes encoded as -1. To fit the framework to different platforms that may not cover all the 450K CpG sites, the undetected features were encoded to 0.

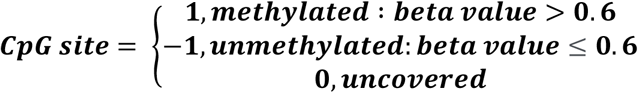

### Model training

The neural network model was trained using 2,801 reference methylomes^8^ generated using Infinium 450K microarrays (Illumina). After binarization of the beta values with threshold 0.6^23^, features with zero variance were filtered, leading to 366,263 CpG sites retained.

To enable the model to take full use of all the information in the features, we sampled the features with a fixed sample rate. During model training, in every iteration, samples in the training set will be randomly masked with the mask rate p, where the masked features will be encoded as 0. To discover the optimal sample rate, we searched and compared different sample rates via 5-fold cross-validation. Finally, sample rate p = 0.25% was selected.

A normalization function and a SoftMax layer was employed to transform the outputs of the neural network into the probabilities of the subtypes of brain tumors. The Adam Optimization Algorithm was used for training. The model was developed and implemented using PyTorch 1.13.0^36^.

### Other analysis

The visualization of genomic information was generated by R package Gviz^37^. Python package seaborn and *PyComplexHeatmap* were used for plotting heatmaps^38^. CpG sites and genes were annotated using Python package CpGtools^39^.

### Data and code availability

Targeted methyl-seq raw data have been deposited at the European Genome-phenome archive (EGA) under accession no. EGAS50000000051. Microarray raw data are provided upon reasonable request. The data and code to recreate all results in this study will be made available upon publication at https://gitlab.com/euskirchen-lab/crossnn. The nanoDx analysis pipeline implementing the crossNN model for end-to-end analysis of nanopore sequencing data is available at https://gitlab.com/pesk/nanoDx. A user-friendly graphical user interface (https://crossnn.dkfz.de) allows to make predictions from methylomes uploaded as bedMethyl files from various platforms and process methylation microarray IDAT files in realtime.

## Data Availability

All data produced in the present study are available upon reasonable request to the authors

